# Zika Virus Congenital Microcephaly Severity Classification and the Association of Severity with Neuropsychomotor Development

**DOI:** 10.1101/2020.07.08.20149120

**Authors:** Nathalia Bianchini Esper, Alexandre Rosa Franco, Ricardo Bernardi Soder, Rodrigo Cerqueira Bomfim, Magda Lahorgue Nunes, Graciane Radaelli, Katherine Bianchini Esper, Aline Kotoski, Willian Pripp, Felipe Kalil Neto, Luciana Schermann Azambuja, Nathália Alves Mathias, Danielle Irigoyen da Costa, Mirna Wetters Portuguez, Jaderson Costa da Costa, Augusto Buchweitz

**Affiliations:** PUCRS, Brain Institute of Rio Grande do Sul (BraIns), Porto Alegre, Brazil; PUCRS, School of Medicine, Neurosciences, Porto Alegre, Brazil; Center for the Developing Brain, Child Mind Institute, New York, New York, USA; Center for Biomedical Imaging and Neuromodulation, Nathan Kline Institute for Psychiatric Research, Orangeburg, New York, USA; Department of Psychiatry, NYU Grossman School of Medicine, New York, NY, USA; Arthur Ramos Memorial Hospital, Imaging Diagnostics – DIRAD, Maceió, Brazil; PUCRS, School of Technology, Porto Alegre, Brazil; PUCRS, Graduate Program in Pediatrics, School of Medicine, Porto Alegre, Brazil; PUCRS, School of Life and Health Sciences, Porto Alegre, Brazil

**Keywords:** brain, congenital microcephaly, development, infants, magnetic resonance imaging, zika virus

## Abstract

**Background:** Zika virus infection during pregnancy is linked to birth defects, most notably, microcephaly, which in its turn, is associated with neurodevelopmental delays.

**Objective:** The goal of the study is to propose a method for severity classification of congenital microcephaly based on neuroradiological findings of MRI scans, and to investigate the association of severity with neuropsychomotor developmental scores. We also propose a semi-automated method for MRI-based severity classification of microcephaly.

**Methods:** Cross-sectional investigation of 42 infants born with congenital Zika infection. Bayley-III developmental evaluations and MRI scans were carried out at ages 13-39 months (mean: 24.8, SD: 5.8). The severity score was generated based on neuroradiologist evaluations of brain malformations. Next, we established a distribution of Zika virus-microcephaly severity score into mild, moderate, and severe and investigated the association of severity with neuropsychomotor developmental scores. Finally, we propose a simplified semi-automated procedure for estimating the severity score, based only on volumetric measures.

**Results:** Results showed a correlation of r = 0.89 (p < 0.001) between the Zika virus-microcephaly severity score and the semi-automated method. The trimester of infection did not correlate with the semi-automated method. Neuropsychomotor development correlated with the severity classification based on radiological readings and with the semi-automated method; the more severe the imaging scores, the lower neuropsychomotor developmental scores.

**Conclusion:** The severity classification methods may be used to evaluate severity of microcephaly and possible association with developmental consequences. The semi-automated methods thus may be an alternative for prediction of severity of microcephaly using only one MRI sequence.

## INTRODUCTION

In 2015, an epidemic of Zika virus infection affected Brazil, especially the northeastern region of the country. Between March 2015 and February 2016, there was a 20-fold increase in births with microcephaly in Brazil, for an equivalent time period [1]. Newborns of mothers infected with Zika virus during pregnancy presented severe brain malformations and abnormalities of development [2], notably, microcephaly [3, 4]. The identification of Zika virus ribonucleic acid (RNA) in the amniotic fluid of mothers whose fetuses had cerebral abnormalities suggested Zika virus transmission occurred during pregnancy [5, 6]. Zika virus-related microcephaly is associated with central nervous system lesions that include destructive, calcification, hypoplasia, and migration disturbances [7]. Moreover, studies show that the brain is susceptible to the effects of Zika virus infection in multiple developmental stages [8].

The developmental outcomes associated with microcephaly (e.g., non-Zika virus-related) may vary. One study showed 50% of children with mild microcephaly had normal intelligence scores [9]. The severity of Zika virus-related microcephaly, however, has been associated with severity of cognitive [10] and motor [11] impairments. In this sense, a combination of developmental and brain imaging evaluations may help predict and better characterize microcephaly-related outcomes, especially in the more recent Zika virus-related microcephaly; a system of evaluation and scoring may help understand and evaluate, with reproducibility, Zika virus-related microcephaly effects and allow for comparison to other cases of microcephaly.

The goal of the present study was to produce a score for the severity of microcephaly using a combination of clinical and brain imaging indices. Previous studies have investigated the relationship between brain imaging of Zika virus-related microcephaly and development [12]. But, to our knowledge, no studies have proposed a brain-imaging based classification of Zika virus-related microcephaly. We further investigated the image characteristics that were more relevant for the classification, and we established a semi-automatic algorithm to predict the severity score, using only the T1-weighted sequence. Lastly, we investigated the relation of the severity score with motor, language, and cognitive development evaluations.

## MATERIALS AND METHODS

### Participants

The study included 42 infants born with suspected or confirmed congenital Zika virus (19 females, mean age: 24.8 months, standard deviation: 5.8 months, head circumference equal or below to 32 cm) registered at a state Health Department. Inclusion criteria for mothers and infants followed Brazilian Ministry of Health guidelines [13]. The criteria for inclusion of the mothers were history of pruritic maculopapular exanthema, positive Zika virus Immunoglobulin M serological reaction, and at least two Zika virus infection symptoms. Inclusion criteria for infants were positive for Zika virus Immunoglobulin G and born to mothers with suspected or confirmed Zika virus infection during pregnancy. Mothers and infants were screened for the following congenital infections disorders, which represented exclusion criteria: syphilis, toxoplasmosis, rubella, cytomegalovirus, and herpes simplex. No infant tested positive for any of these infections. Written informed consent was obtained from the parents or guardians of the infants. The study was approved by the research ethics committee of the Pontifical Catholic University of Rio Grande do Sul (CAEE 61642016.6.1001.5336).

### Instruments and procedures

Bayley Scales of Infant and Toddler Development (Bayley-III) scores were collected one day after the MRI exam. We used the Brazilian Portuguese version of the Bayley-III [14] scales to assess three domains: cognition, language (receptive and expressive communication), and motor (gross and fine). HC was measured at birth and at the MRI exam; we also generated a HC growth ratio score by subtracting HC at birth from HC on the day of MRI exam and dividing the result by HC at birth.

### MRI data acquisition

Twenty-eight infants were scanned at Brain Institute of Rio Grande do Sul in Porto Alegre, Rio Grande do Sul and 14 were scanned at the Memorial Hospital Arthur Ramos in Maceió, Alagoas; see the online supplementary material 1 for a detailed description of the sequence parameters. To avoid head motion artifacts during MRI scans, participants in both sites were anesthetized. The MRI exams were carried out under the supervision of an expert anesthesiologist.

### Zika virus-microcephaly severity score

Brain MRI images were analyzed by two neuroradiologists (R.B.S – 15 years of experience; R.C.B – 15 years of experience). They performed independent analysis of the MRI images. The neuroradiologists were aware they were evaluating Zika virus-related microcephaly cases, but they were blind to medical history or clinical information about the patient; they were also blind to each other’s evaluation. After the neuroradiological evaluations were performed, the disagreements in evaluations were resolved by means of reaching a consensus in a case-by-case discussion.

Brain MRI images were reviewed to investigate structural abnormalities based on 13 characteristics. The imaging characteristics were drawn from studies of brain effects of congenital microcephaly in babies born during the Zika virus outbreak in Brazil [8, 15–17]. Except for calcification, the characteristics were scored on a 4-point scale from zero to 3, zero represented normality and 3 the most severe abnormality, according to the neuroradiologist. The brain calcifications, in turn, were scored on a 5-point scale from zero to 4, zero represented absence of calcifications, and 4 the presence of calcifications in 4 or more regions. The brain regions analyzed for the calcifications were cortico-subcortical white matter junction, periventricular, basal ganglia, and posterior fossa. We used five imaging sequences to evaluate the imaging characteristics. The specific imaging sequence employed for each score is described in online supplementary material 2. There were disagreements between two neuroradiologists in 24 of 546 characteristics.

The Zika virus-microcephaly severity score grouping was established using the scores of the 13 malformations of the different MRI structural scans. The three categories were established dividing the total score in terciles: severity scores ranging from 0 to 12 were classified as mild microcephaly; from 12 to 25, as moderate microcephaly; and, finally, scores above 26 were classified as severe microcephaly. The maximum score is 41.

#### Semi-automated version using T1-weighted MRI-imaging only

We present a method to semi-automatically create the Zika virus-microcephaly severity score and establish severity of microcephaly-related abnormalities. The motivation to create this semi-automated method was to remove the subjectivity of radiological findings, and thus establish reproducibility for the severity score. The method is based on MRI volumetric measures alone. The goal was to predict the severity score using only one brain image sequence (T1 structural scan) that delineates four volumes of interest (VOI) for each participant. The volumes were: (a) the lateral ventricles, (b) whole brain, (c) intracranial, and (d) the cerebellum segmentation (see online supplementary material 3). We applied a semi-automated region growing segmentation method based on the edge detection algorithm in the Insight Segmentation and Registration Toolkit software [18]. The regression model and the procedure for segmentation of the VOIs are described in online supplementary material 4. This procedure allows for replication of evaluation across sites and research of microcephaly-related developmental outcomes independent of neuroradiological evaluations. The goal, of course, is not to replace neuroradiological evaluation but rather to afford an instrument for research purposes.

### Statistical Analysis

We carried out statistical analyses using the Statistical Package for Social Sciences (IBM SPSS Statistics, RRID:SCR_019096), version 23. We used descriptive statistics for the population demographics. We used Kolmogorov-Smirnov nonparametric test to evaluate whether clinical variables (age, head circumference at birth and on day of MRI exam, and trimester of infection) and volumetric variables had a normal distribution. The Kolmogorov-Smirnov test showed that all variables had a non-parametric distribution. Next, we applied Spearman correlation to investigate the relationship among variables. We used partial correlations and controlled for age for the analyses of Bayley-III scales and the clinical and image variables. We used the raw score of Bayley-III to perform analysis of correlations among cognitive, receptive and expressive communication, and fine and gross motor scores with the imaging readings scores. The alpha coefficient of Cronbach was calculated to verify the internal consistency, measuring the reliability of the radiological severity score. We performed a linear regression analysis to create the semi-automated Zika virus-microcephaly severity Score (dependent variable) by using the intracranial volume (*x*_1_), ratio between lateral ventricles and brain (*x*_2_), ratio between brain and intracranial (*x*_3_), and the square of each one of the three volumetric variables 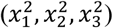. A p < 0.05 was considered statistically significant for all analyses.

## RESULTS

### Participant’s demographics

We evaluated 42 infants with confirmed Zika virus infection during pregnancy. The participants mean birth weight was 2,650g (SD: 0.530g); 37 participants were born to full term. The mean head circumference (HC) at birth was 29.81 cm (SD: 1.89 cm). Nine infants were born with HC < 1SD, ten with HC < 2SD and twenty-three with HC < 3SD below the mean for gestational age. Head circumference was also measured on the day of the MRI exam, where one infant presented a normal HC, three with HC < 2SD, twenty-six with HC < 3SD and twelve without the measure of HC. Timing of maternal infection was self-reported, according to the trimester of pregnancy that mothers had Zika virus symptoms. Twenty-one mothers reported that the timing of infection occurred in the first trimester of pregnancy, twelve, in the second trimester, five, in the third trimester and four mothers could not recall the trimester of infection.

#### Zika virus-microcephaly severity classification

Brain malformations varied considerably across participants (please see online supplementary material 5 for brain images for all participants). The observed variation corroborates previous studies about Zika virus-related microcephaly malformations [19, 20]. Seven participants were classified in the mild range (16.7%), 13 participants in the moderate range (31%) and 22 in the severe range (52.4%). We obtained an alpha of Cronbach = 0.937, which means that the severity classification items have high consistency. Table 1 presents the image characteristics evaluated, their classifications, and the intraclass correlation coefficient. Online supplementary material 6 shows examples of the mild, moderate, and severe categories of each imaging characteristics.

**Table 1.** Description of the image interpretation for the Zika virus-microcephaly severity score.

| Brain Image Characteristic | 0 <sup>a</sup> | Score |  |  |  | ICC |
| --- | --- | --- | --- | --- | --- | --- |
|  |  | 1 | 2 | 3 | 4 |  |
| Cephalic Perimeter Reduction |  | Mild | Moderate | Severe | -- | 0.977 |
| Volume Reduction |  | Mild | Moderate | Severe | -- | 0.970 |
| Enlarged Supratentorial Subarachnoid Space |  | Mild | Moderate | Severe | -- | 0.968 |
| Ventriculomegaly |  | Mild | Moderate | Severe | -- | 1 |
| White Matter Volume Reduction |  | Mild | Moderate | Severe | -- | 1 |
| Myelination (hypomyelination/demyelination) |  | Mild | Moderate | Severe | -- | 1 |
| Gyral Pattern Simplification (*n* of lobes) |  | Focal (1) | Moderate (2) | Diffuse (3+) | -- | 1 |
| Hippocampus |  | Malrotation | Volume reduction | Malrotation + volume reduction | -- | 1 |
| Corpus Calossum hypoplasia/dysgenesis |  | Mild | Moderate | Severe | -- | 0.991 |
| Brain Calcifications (*n* of regions) <sup>b</sup> |  | 1 | 2 | 3 | 4 + | 0.967 |
| Brainstem hypoplasia |  | Mild | Moderate | Severe | -- | 0.899 |
| Cerebellar Volume hypoplasia |  | Mild | Moderate | Severe/agenesis | -- | 0.987 |
| Malformations of Cortical Development: polymicrogyria/focal pachygyria (*n* of lobes) |  | Mild (1) | Moderate (2) | Diffuse (3 or +) | -- | 0.993 |

<sup>a</sup> 0 = Normal. <sup>b</sup> Number of regions: the location of brain calcification evaluated was cortico-subcortical white matter junction, periventricular, basal ganglia, and posterior fossa. ICC = Intraclass Correlation Coefficient between two radiologists for each image interpretation score. -- = score does not apply.

#### Radiological interpretation

Results showed a correlation among cephalic perimeter reduction and all image characteristics that make up the Zika virus-microcephaly severity score; the association among cephalic perimeter reduction and the imaging indices suggests Zika virus has a generalized effect on the brain. Figure 1 shows sagittal MRI images for nine subjects; the images are rank-ordered from lowest to highest Zika virus-microcephaly severity scores to illustrate the brain abnormalities characteristic of each severity group.

**Figure 1.**
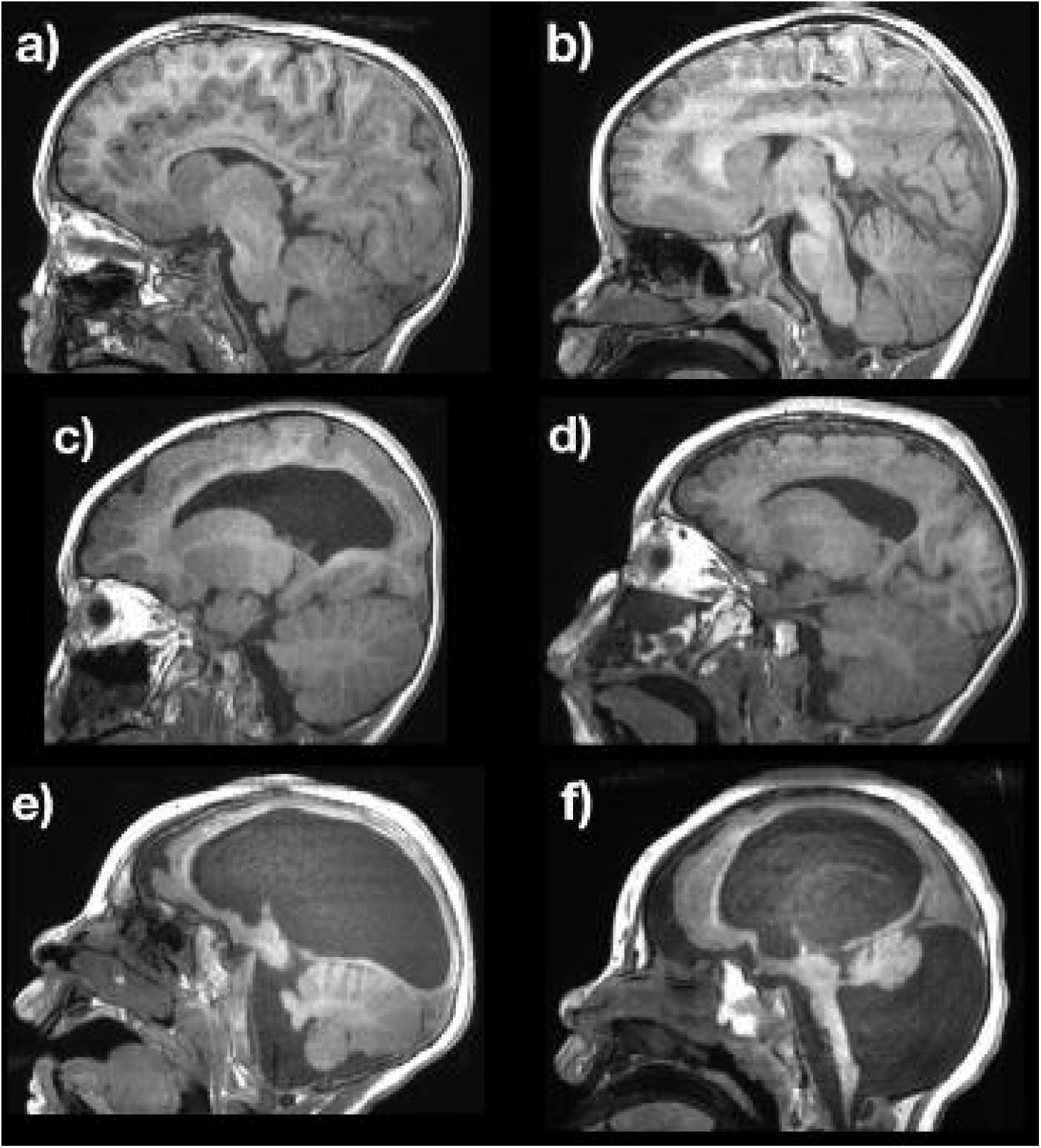
Images were rank-ordered from lowest to highest Zika virus-microcephaly severity score, age range 22-30 months old. In the mild group: (a) boy, head circumference at birth (HCb) = 32 cm (< 1SD), head circumference at MRI exam (HC) = 46 cm (< 2SD), Zika virus-microcephaly severity score = 5, and semi-automated score = 3; and (b) a girl, HCb = 32 cm (< 1SD), HC = 41cm (< 3SD), zika virus-microcephaly severity score = 6, and semi-automated score = 6. In the moderate group: (c) a boy, HCb = 31cm (< 2SD), HC = 38cm (< 3SD), zika virus-microcephaly severity score = 17, and semi-automated score = 17; and (d) a boy, HCb = 31cm (< 2SD), HC = 42cm (< 3SD), zika virus-microcephaly severity score = 22, and semi-automated score = 18. In the severe group: (e) a girl, HCb = 27cm (< 3SD), HC = 40cm (< 3SD), zika virus-microcephaly severity score = 33, and semi-automated score = 32; and (f) a girl, HCb = 25cm (< 3SD), HC = 39cm (< 3SD), zika virus-microcephaly severity score = 38, and semi-automated score = 36. SD = standard deviation; HCb = head circumference at birth; HC – head circumference at MRI exam. Head circumference was expressed in centimeters and normalized by Z-score. Sagittal T1w MR images illustrate differences in brain morphology among participants in the three severity groups.

The correlation between all image characteristics was performed and presented in Figure 2. All correlations showed to be statistically significant (r ≥ 0.32, p < 0.05). Individual Zika virus-microcephaly severity scores per participant are presented in online supplementary material 7. The correlations among the trimester of Zika virus infection and brain abnormalities that make up the Zika virus-microcephaly severity score were calculated. Results indicated that trimester of infection was significantly correlated with cephalic perimeter reduction (r=-0.334, p=0.04), cortical development (r = -0.655, p < 0.001), gyral simplification (r = -0.654, p < 0.001), and calcification (r = -0.347, p = 0.033).

**Figure 2.**
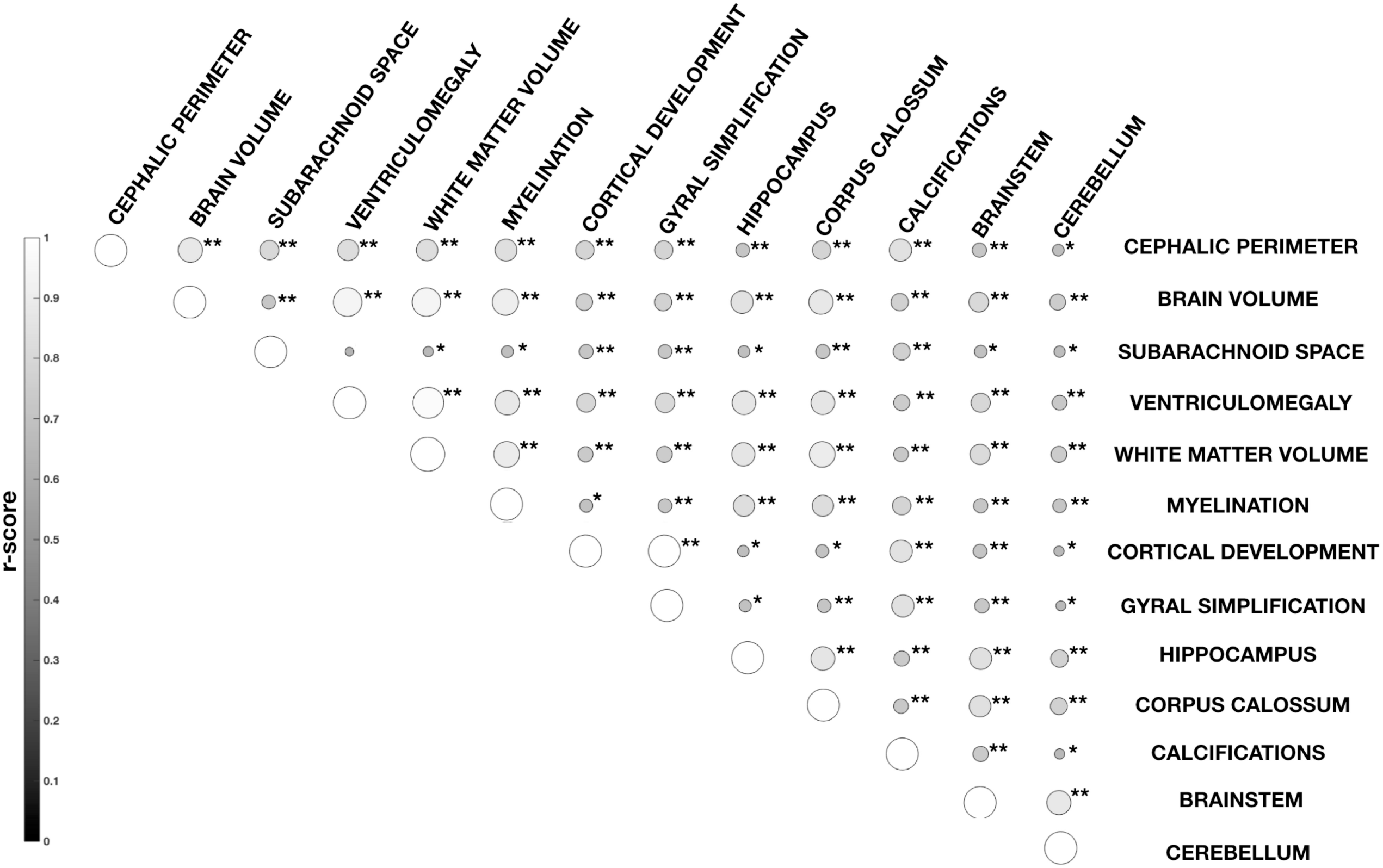
Correlation matrix among the 13 imaging characteristics evaluated on all 42 participants. Lighter shades of gray and larger circles represent higher correlation (r-score). Statistical significance is identified by * p < 0.01 and ** p < 0.001.

#### Neuropsychomotor development and brain imaging indices

The neuropsychomotor developmental Bayley-III scores were all found below average for participant age. Our results show a significant correlation between the Zika virus-microcephaly severity score and the Bayley-III scales domains. Bayley-III scores correlated only with head circumference at birth, in all domains: cognitive (r = 0.373; p = 0.018), receptive (r = 0.369; p = 0.019) and expressive communication (r = 0.333; p = 0.036), fine (r = 0.46; p = 0.003) and gross motor (r = 0.423; 0 = 0.007). There was no significant correlation between Bayley-III scores and HC scores at the day of the MRI exam or Bayley-III scores and HC growth ratio. Neither the trimester of infection or the HC growth ratio at the time of developmental evaluation were significantly associated with cognitive, language and motor scores. Correlation results are presented in the online supplementary material 8.

Results showed an association among the imaging characteristics that composes the Zika virus-microcephaly severity scores, and neuropsychomotor developmental outcomes. The association involved fine and gross motor, receptive and expressive communication, and cognitive evaluations. On further analyses, increased severity in the posterior fossa (including reduction of cerebellar volume and brainstem) had a correlation with motor skills. Previous studies have shown that alterations in the posterior fossa can cause devastating balance and motor problems [21]. The correlation results are presented in Table 2.

**Table 2.** Correlation results among Bayley-III scales and the imaging characteristics of Zika virus-microcephaly severity score.

|  | **Cognition**<br>**r (p-value)** | **Receptive**<br>**Communication**<br>**r (p-value)** | **Expressive**<br>**Communication**<br>**r (p-value)** | **Fine Motor**<br>**r (p-value)** | **Gross Motor**<br>**r (p-value)** |
| --- | --- | --- | --- | --- | --- |
| **Cephalic Perimeter** | -0.51* | -0.44* | -0.41* | -0.56** | -0.45* |
| **Brain Volume** | -0.59** | -0.59** | -0.55** | -0.67** | -0.61* |
| **Subarachnoid Space** | -0.31* | -0.43* | -0.39* | -0.38* | -0.41* |
| **Ventriculomegaly** | -0.56** | -0.56** | -0.51* | -0.64** | -0.56** |
| **White Matter**<br>**Volume** | -0.63** | -0.62** | -0.5* | -0.68** | -0.58** |
| **Myelination** | -0.53** | -0.45* | -0.36* | -0.51* | -0.31 |
| **Gyral Pattern**<br>**Simplification** | -0.37* | -0.38* | -0.41* | -0.46* | -0.37* |
| **Hippocampus** | -0.38* | -0.39* | -0.29 | -0.45* | -0.39* |
| **Corpus Callosum** | -0.53** | -0.54** | -0.52* | -0.61** | -0.6** |
| **Brain Calcifications** | -0.28 | -0.19 | -0.19 | -0.31* | -0.24 |
| **Brainstem** | -0.33* | -0.27 | -0.28 | -0.42* | -0.4* |
| **Cerebellar Volume** | -0.3 | -0.31* | -0.31* | -0.34* | -0.31* |
| **Malformations of**<br>**Cortical Development** | -0.36* | -0.37* | -0.41* | -0.45* | -0.37* |

2-tailed partial correlation, controlling by age (in months); \* $p < 0.05$ ; \*\* $p < 0.001$

**Table 3.** Correlation results among the 75% random sample and the neuropsychomotor development.

|  | r | p | Standard error | 95% CI |
| --- | --- | --- | --- | --- |
| Zika virus-microcephaly severity score | 0.878 | <0.001 | 0.06 | 0.716; 0.950 |
| Semi-automated severity score | 0.984 | <0.001 | 0.01 | 0.930; 0.998 |
| HC at birth | -0.739 | <0.001 | 0.08 | -0.864; -0.529 |
| HC at MRI exam | -0.380 | 0.038 | 0.18 | -0.686; -0.02 |
| HC growth ratio | 0.187 | 0.324 | 0.201 | -0.234; 0.537 |
| Trimester of infection | -0.086 | 0.607 | 0.162 | -0.392; 0.233 |
| Cognition<sup>a</sup> | -0.619 | <0.001 | 0.136 | -0.777; -0.219 |
| Receptive communication<sup>a</sup> | -0.595 | <0.001 | 0.142 | -0.755; -0.215 |
| Expressive communication<sup>a</sup> | -0.542 | <0.001 | 0.164 | -0.748; -0.107 |
| Fine motor<sup>a</sup> | -0.713 | <0.001 | 0.114 | -0.843; -0.398 |
| Gross motor<sup>a</sup> | -0.622 | <0.001 | 0.149 | -0.795; -0.190 |

<sup>a</sup>partial correlation, controlling by age; assessed through Bayley-III scales.

#### Semi-automated version of the Zika virus-microcephaly severity score

We created a semi-automated method to simplify the generation of the Zika virus-microcephaly severity score, using brain-based volumes and one brain image sequence only (T1-weighted volumetric image). We used a linear regression to predict the severity score (dependent variable) by using the measured brain volumes as independent variables. The β values obtained through linear regression are the following: β_0_ = 51.404, β_1_ = -12.665, β_2_ = -4.829, β_3_ = -10.789, β_4_ = 1.28, β_5_ = 1.311 and β_6_ = -28.327. The scatter plot of the linear regression is shown in Figure 3.

**Figure 3.**
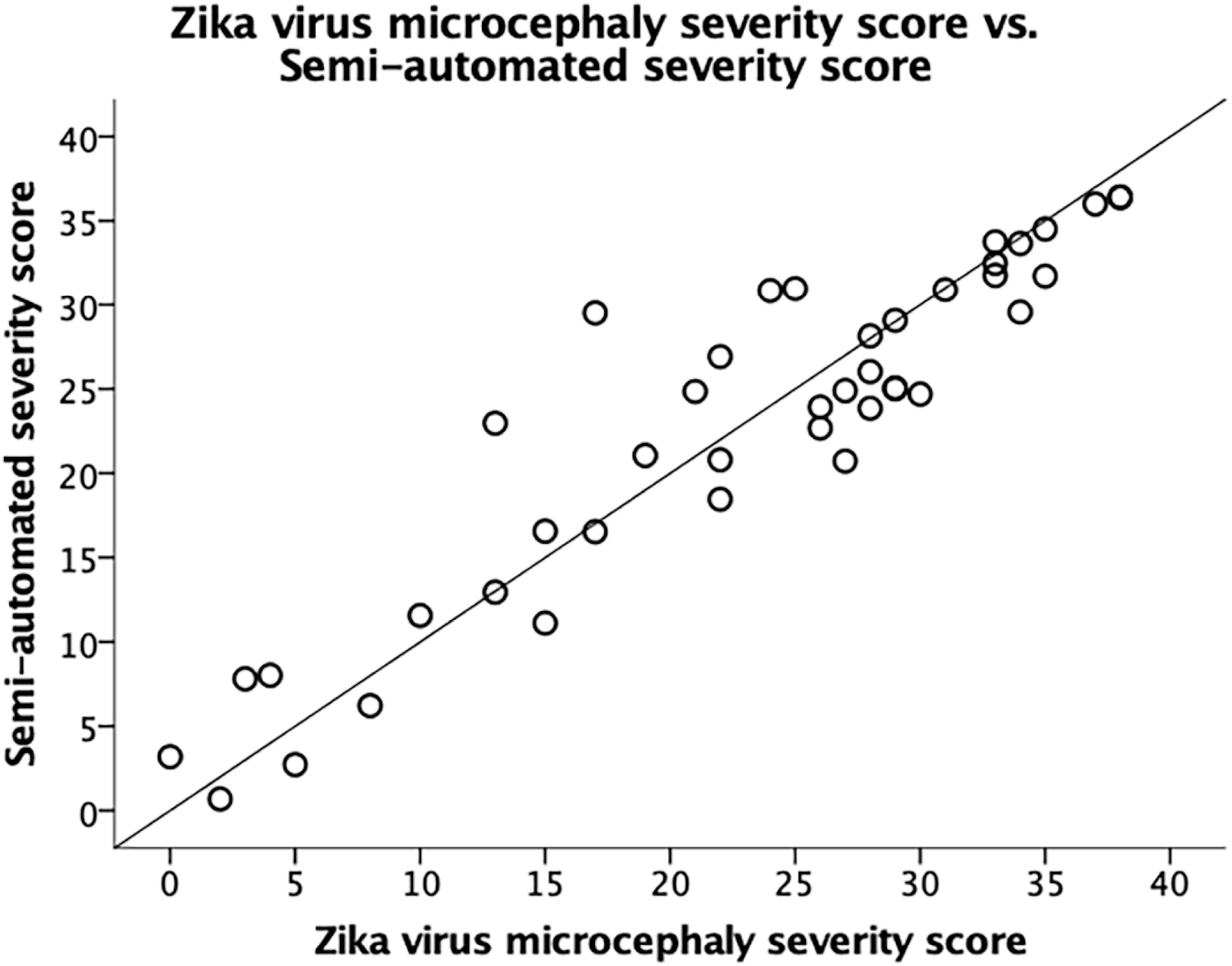
Zika virus-microcephaly severity score (x axis) versus semi-automated severity score (y axis). The dashed line represents y = x. This linear behavior indicates a strong correlation between the two score (r = 0.89; p < 0.001; 2-tailed; Spearman).

We also used the semi-automated score to establish the proposed classification of severity into groups. Using terciles, the scores for the mild group ranged from 0 to 13 and from 14 to 27 for the moderate group. Participants with the predicted severity score above 27 were classified into the severe group. Considering the predicted scores, nine participants were classified in the mild group (21.4%), 16 participants in the moderate group (38.1%) and 17 participants in the severe group (40.5%). Grouping for each participant can be seen in online supplementary material 9.

We validated the semi-automated score by performing a bootstrap analysis (1,000 bootstrap samples) in a 75% random sample (32 participants). Now, the β values obtained through linear regression in the random sample are: β_0_ = 54.774, β_1_ = -1.836, β_2_ = -5.240, β_3_ = -27.869, β_4_ = -9.666, β_5_ = 1.454 and β_6_ = -14.326. Table 4 shows the correlation results among the 75% random sample, trimester of infection, HC, and the neuropsychomotor development assessed thought Bayley-III.

**Table 4.** Comparison between the Zika virus-microcephaly severity score and the semi-automated severity score

|  | Zika virus-microcephaly<br>severity score | Semi-automated<br>severity score |
| --- | --- | --- |
| **Score ranges** |  |  |
| Mild | 0 – 12 | 0 – 12 |
| Moderate | 13 – 25 | 13 – 25 |
| Severe | 26 – 38 | 26 – 36 |
| **Participants per group - N. (%)** |  |  |
| Mild | 7 (16.7%) | 9 (21.4%) |
| Moderate | 13 (31%) | 17 (40.5%) |
| Severe | 22 (52.4%) | 16 (38.1%) |
| **Correlation analysis with score - r (p-value)** |  |  |
| Trimester of infection | -0.37* | -0.1 (p = 0.5) |
| HC at birth | 0.65** | -0.67** |
| HC at MRI exam | -0.35 (p = 0.054) | -0.3 (p = 0.08) |
| HC growth ratio | 0.15 (p = 0.4) | 0.2 (p = 0.1) |
| Cognition <sup>a</sup> | -0.58** | -0.6** |
| Receptive Communication <sup>a</sup> | -0.56** | -0.59** |
| Expressive Communication <sup>a</sup> | -0.52** | -0.54** |
| Fine Motor <sup>a</sup> | -0.66** | -0.7** |
| Gross Motor <sup>a</sup> | -0.57** | -0.62** |

\* p < 0.05; \*\* p < 0.001; <sup>a</sup> from the Bayley-III assessment; 2-tailed Spearman correlation. Partial correlation used for correlations with Bayley-III scales, controlling for age.

### Comparisons between severity scores and development

Table 4 presents a set of results for the two versions of the severity score: the Zika virus-microcephaly severity score based on 13 image characteristics, and the semi-automated severity score. The trimester of infection correlated only with Zika virus-microcephaly severity score; the trimester is an estimate that is error-prone, and possibly not granular and accurate enough to provide information about severity of microcephaly. However, the head circumference at birth correlated with both severity scores. Head circumference measure at the MRI exam and growth ratio did not significantly correlate with any of the severity scores.

Regarding to the neurodevelopment, the severity scores significantly correlated with the neuropsychomotor development assessed through the Bayley-III scales. The correlation was greater for the semi-automated than for the radiological-based scores. The stronger correlations suggest the semi-automated score provides an assessment of severity that may be informative of the clinical stage of neuropsychomotor development.

## DISCUSSION

To our knowledge, this is the first study to propose a brain malformation severity classification for Zika virus-related microcephaly based on radiological findings. The severity of microcephaly was associated with poorer developmental scores in all cognitive domains. Head circumference is a widely used parameter for assessing severity of microcephaly [20]. Our study shows infants presented significant variation in brain morphological malformations regardless of head circumference measured at the time of the brain scan (or the HC growth ratio). Only HC measured at birth showed association with developmental scores. The Zika virus-microcephaly severity score is a more fine-grained evaluation and significantly correlated with developmental scores. This scoring system may capture the effects of Zika virus on brain development with a granularity that allows for investigating prognoses of cognitive development in longitudinal studies of microcephaly. We also showed that the information of these brains volume can suffice to classify participants into severity groups (mild, moderate, and severe).

### Grouping brain malformations according to severity

Grouping participants according to severity (mild, moderate, and severe) is based on the more granular categorization of the Zika virus-microcephaly severity score. Thus far, this scoring system seems to better inform the probability that Zika virus-related microcephaly will impact cognitive and behavior outcomes: it correlated with all Bayley-II scores. However, the semi-automated score has the potential of being further developed and adjusted by establishing, for example, a weighted score for abnormalities and indices that have a more significantly load on development. The grouping of malformations according to the severity score may be tested in future evaluations against neurodevelopmental outcomes in infants with congenital microcephaly.

### Semi-automated Zika virus-microcephaly severity score

We have shown that there is a significant correlation between the Zika virus-microcephaly severity score and the semi-automated scores based on the VOIs (r = 0.891; p < 0.001). However, there were disagreements between the clinical and the semi-automated scores. Participant ID = Z101 had a Zika virus-microcephaly severity score of 17 and a semi-automated score of 30. These scores categorize microcephaly as moderate according to the radiological finding, but as severe according to the semi-automated method. Visual inspection of the images suggests the semi-automated score is biased by enlarged ventricles and reduced cerebral cortex volume. Again, further development of the score and testing with other microcephalic populations may help adjust, for example, how indices are weighed in the semi-automated. Nonetheless, the semi-automated method for measuring brain abnormalities can inform neuroradiological findings and neurological evaluations. The training required to perform the segmentations requires moderate knowledge about brain anatomy and about use of a software called “Insight Segmentation and Registration Toolkit” (ITK-SNAP; University of Pennsylvania and University of Utah) [18]. It is expected that with training in the procedures the results can be easily replicated across research studies.

### Study limitations

The study limitations include small sample size relative to the number of microcephalic infants born during the epidemic. Evidently, larger and more heterogeneous samples allow for a better evaluation of replicability and generalizability of the system proposed, and of its ability to inform prognoses. We underscore that sample size and heterogeneity was limited the timing of the outbreak, relative to the beginning of the study (the outbreak was mostly contained once we started investigating the effects, which limited the ability to include newborn patients and to observe pregnancies), and by logistical challenges: participants were flown from northeastern regions of the country (more than 3,600 km away from our research site) to participate in the study. Additionally, the study did not have a control group and the method was not applied to either controls or another population of infants with brain abnormalities. Lastly, Zika virus congenital infection may result in neuronal loss, disruption, and destruction; however, our study was limited to the assessment of malformations. Anatomopathological data were not collected in the present study.

## CONCLUSION

This study provides a method for severity classification of brain abnormalities, establishing three categories: mild, moderate, and severe microcephaly. This severity classification relies on assessment of combination of alterations in brain structures and indices; is thus more granular and possibly more promising for understanding patient prognoses relative to classification of severity based on head circumference alone. The classification system must be further tested and evaluated. The proposed classification may apply to other populations with microcephaly (not related to Zika virus) or with other congenital, brain-related diseases.

## Data Availability

Technical and statistical code are available from the first author at

