## Supplementary material for "Zika Virus Congenital Microcephaly Severity Classification and the Association of Severity with Neuropsychomotor Development": Online supplementary material 1

Brain Institute: The data was collected using a GE HDxT 3T MRI scanner with an 8-channel head coil. T1 structural scans were acquired with the following parameters using a BRAVO sequence: repetition time (TR) = 6.16ms, echo time (TE) = 2.18ms, flip angle = 11°, acquisition matrix of 240 x 240 x 196 and voxel size of 1.0 x 1.0 x 1.0mm.

Memorial Hospital: The data was collected on an Optima MR450W 1.5T MRI scanner with a 16-channel head coil. The T1 structural scans were acquired with the following parameters using a BRAVO sequence: TR = 8.7ms, TE = 3.224ms, flip angle = 12°, acquisition matrix = 256 x 256 x 100 and voxel size = 0.938 x 0.938 x 1.2mm.
