## Supplementary material for "Zika Virus Congenital Microcephaly Severity Classification and the Association of Severity with Neuropsychomotor Development": Online supplementary material 2

MRI Sequences and Image Evaluation

| Brain Imaging Characteristic | MRI Sequence |
| --- | --- |
| Cephalic Perimeter Reduction | 3D FSPGR |
| Volume Reduction | 3D FSPGR |
| Enlarged Supratentorial Subarachnoid Space | Axial/Coronal/Sagittal T2 FSE |
| Ventriculomegaly | 3D FSPGR  Axial/Coronal/Sagittal T2 FSE |
| White Matter Volume Reduction | 3D FSPGR  Axial/Coronal/Sagittal T2 FSE |
| Myelination | 3D FSPGR  Axial/Coronal/Sagittal T2 FSE |
| Gyral Pattern Simplification | 3D FSPGR  Axial/Coronal/Sagittal T2 FSE |
| Hippocampus | 3D FSPGR  Coronal T2 FSE |
| Corpus Calossum hypoplasia/  dysgenesis | 3D FSPGR |
| Brain Calcifications | Axial T2* SWI |
| Brainstem hypoplasia | 3D FSPGR |
| Cerebellar Volume Hypoplasia | 3D FSPGR  Axial/Coronal/Sagittal T2 FSE |
| Cystic Malformation of Posterior Fossa | 3D FSPGR  Axial/Coronal/Sagittal T2 FSE |
| Malformations of Cortical Development | 3D FSPGR  Axial/Coronal/Sagittal T2 FSE |
