## Supplementary material for "Zika Virus Congenital Microcephaly Severity Classification and the Association of Severity with Neuropsychomotor Development": Online supplementary material 3

Example of the volumes of interest (VOI) after segmentation, for one participant. Orange-colored volumes represent the result of the semi-automated segmentation method.

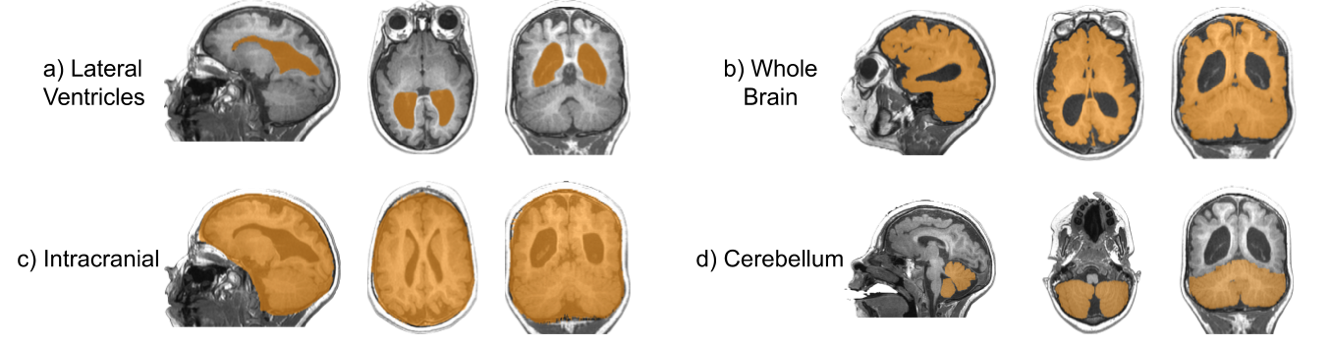
