## Supplementary material for "Zika Virus Congenital Microcephaly Severity Classification and the Association of Severity with Neuropsychomotor Development": Online supplementary material 4

**Regression model**

We performed a linear regression analysis to predict the Zika virus-microcephaly severity score (dependent variable) by using the intracranial volume $(x_{1})$, ratio between lateral ventricles volume and brain volume $(x_{2})$, ratio between brain volume and intracranial volume $(x_{3})$, and the square of each one of the three volumetric variables $(x_{1}^{2},x_{2}^{2},x_{3}^{2})$:

$y= \beta_{0}+\beta_{1}x_{1}+\beta_{2}x_{2}+\beta_{3}x_{3}+\beta_{4}x_{1}^{2}+\beta_{5}x_{2}^{2}+\beta_{6}x_{3}^{2}+e$	(1)

In Equation 1, $y$ is the Zika virus-microcephaly severity score, the β ‘s are the coefficients given by the regression analysis, and $e$ is the residual.

**Volumetric measures – Volume of interest (VOI)**

For each VOI, we added at least two control points inside the region of interest. If necessary, additional control points were added near tissue/region boarder surfaces to improve the accuracy of the segmentation procedure. The size of VOIs were increased until one part of the VOI touched a border surface. For example, when segmenting the lateral ventricles, the border surface area is defined as the surface between the ventricles and white matter. The volume growing procedure underestimates the size of the VOI. Finally, to improve the accuracy of segmentation, we manually adjusted the segmentation though visual inspection by including any missing voxels inside the respective VOI. We calculated the volume of the final VOI (in liters) after segmentation processes were carried out. All 42 T1 images were independently segmented by three collaborators (K.B.E, A.K. and W.P.) and subsequently checked by a fourth researcher (N.B.E.). Inter-rater reliability for the four VOIs showed intraclass correlation coefficient (ICC) as excellent (>0.8) for all regions (ICC scores: ventricles = 0.974, whole brain = 0.9945, intracranial = 0.8939 and cerebellum = 0.9832). The proposed semi-automated score requires about 4 hours of manual labor to calculate the VOI per sample.
