## Supplementary material for "Zika Virus Congenital Microcephaly Severity Classification and the Association of Severity with Neuropsychomotor Development": Online supplementary material 5

Sagittal MRI images for all participants showing significant differences in brain morphology between participants.

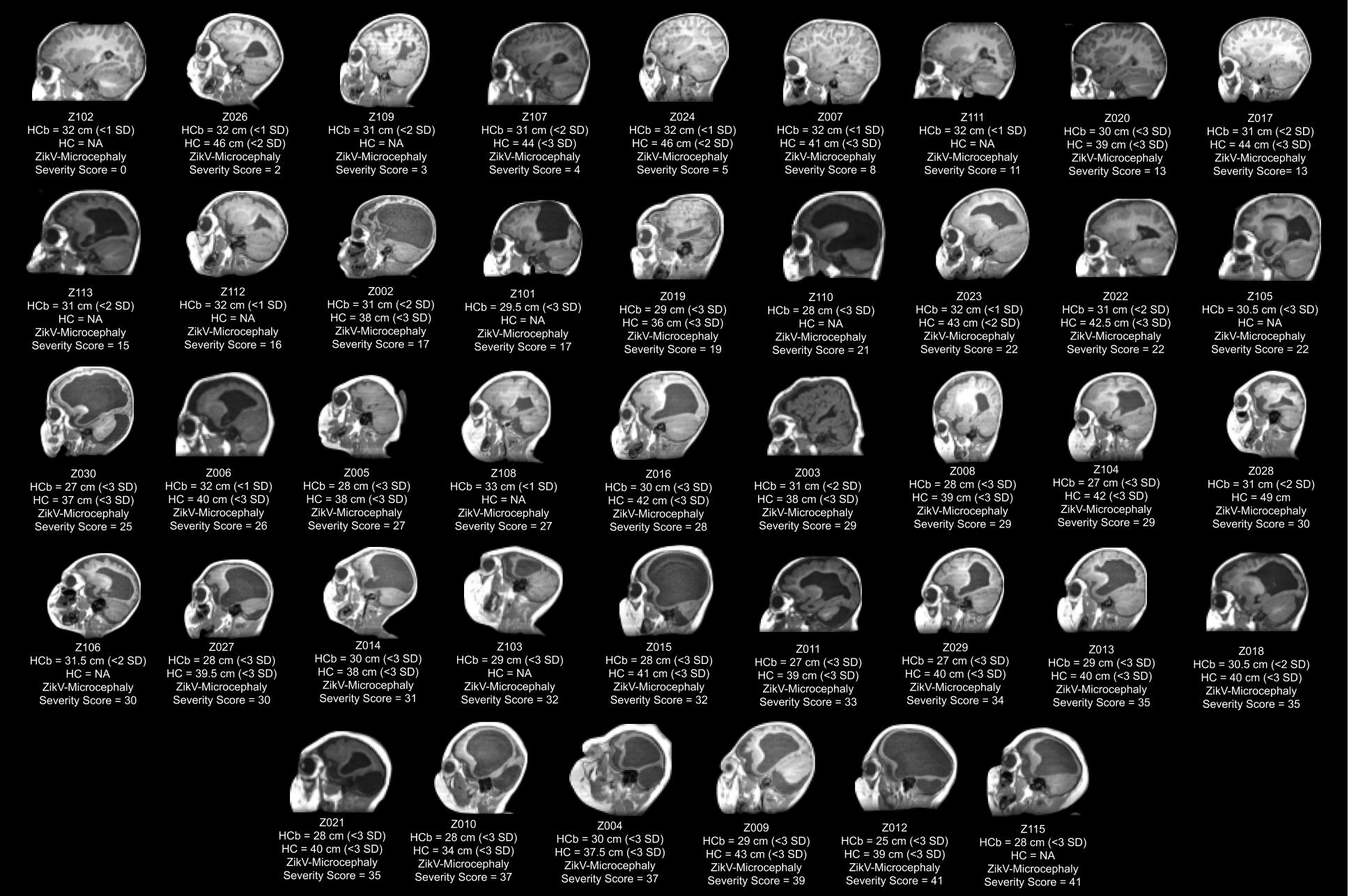

SD = standard deviation; HCb = head circumference at birth; HC = head circumference at MRI exam. Head circumference was expressed in centimeters and normalized by Z-Score. Images were organized by ZikV-Microcephaly Severity Score (simplified version with 6 image items), from lowest to highest.
