## Supplementary figures and images for "Zika Virus Congenital Microcephaly Severity Classification and the Association of Severity with Neuropsychomotor Development"

### Online supplementary material 6

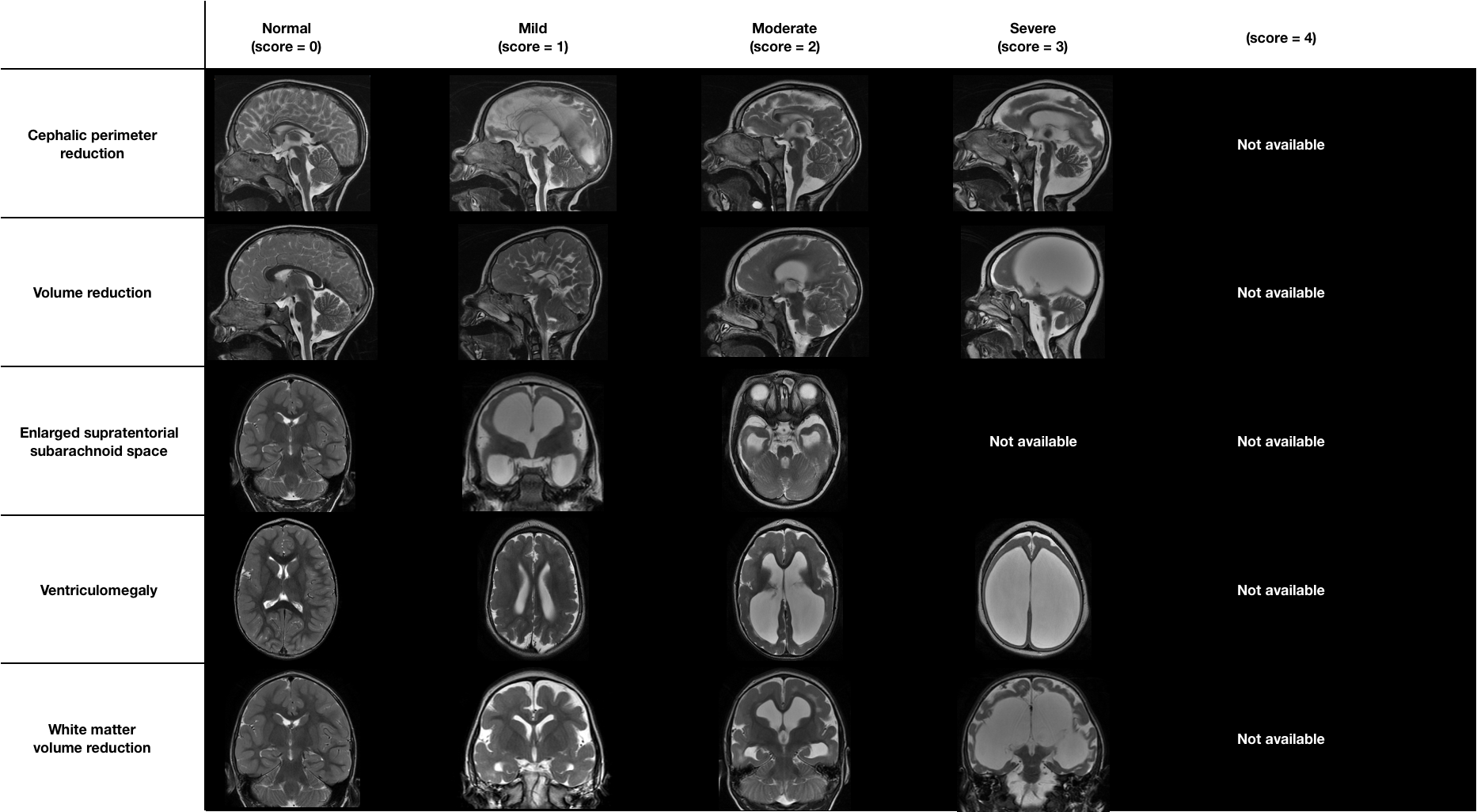

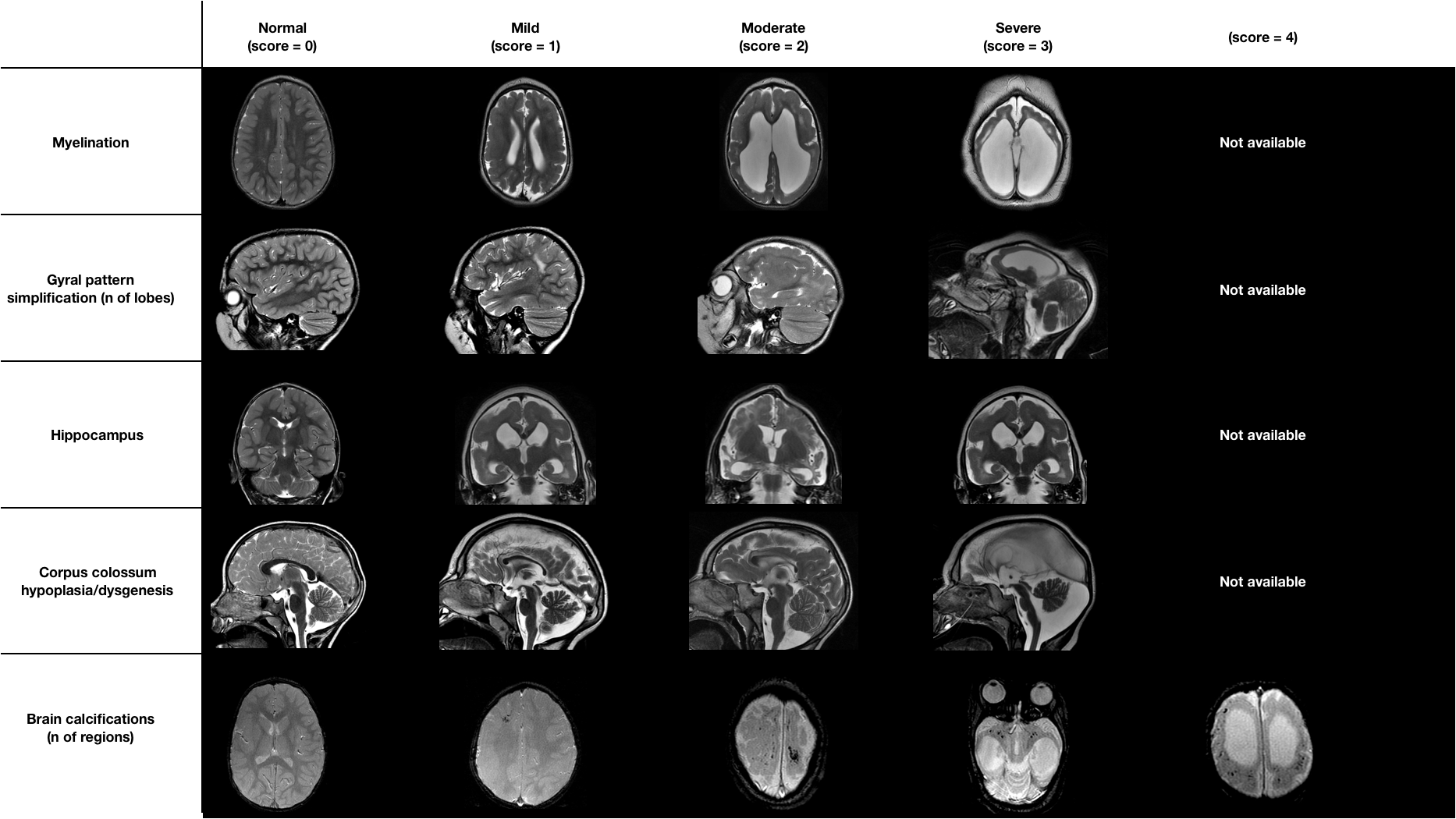

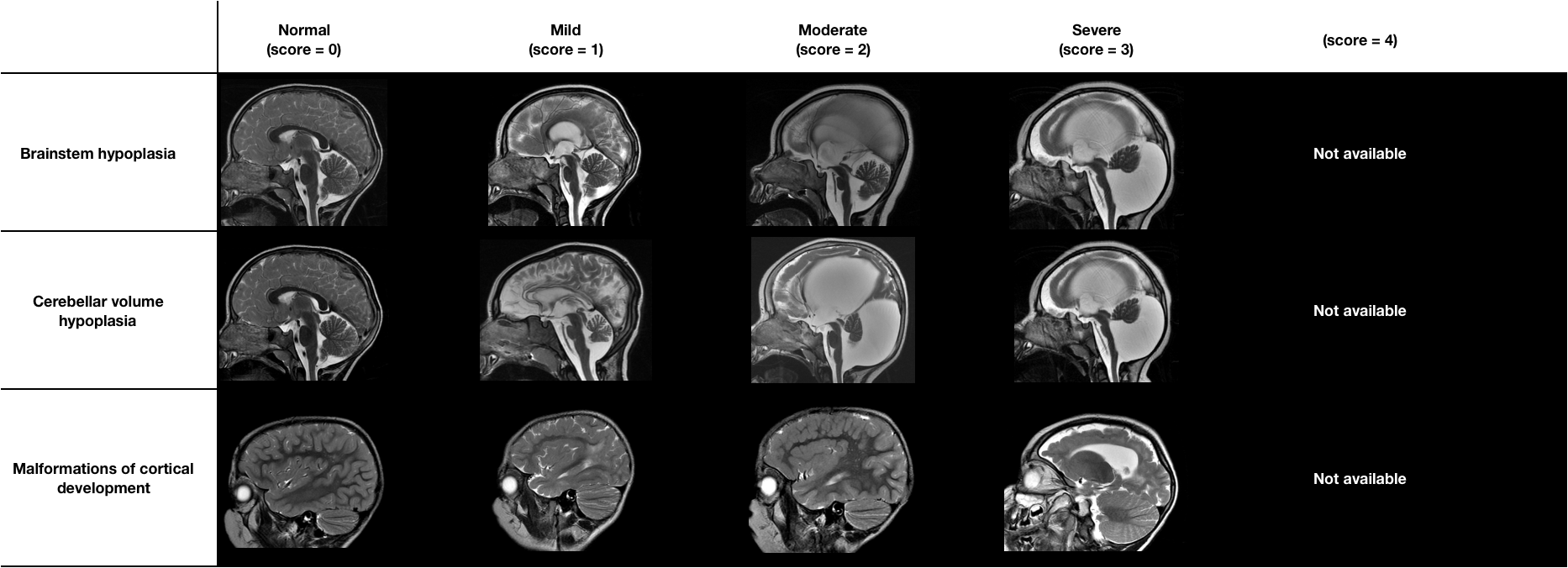
