## Supplementary material for "Zika Virus Congenital Microcephaly Severity Classification and the Association of Severity with Neuropsychomotor Development": Online supplementary material 7

Zika virus-microcephaly severity score indices

| ID | Cephalic Perimeter | Brain Volume | Supratentorial Subarachnoid Space | Ventriculomegaly | White Matter Volume | Myelination | Gyral Pattern Simplification | Hippocampus | Corpus Callosum | Brain Calcifications | Brainstem | Cerebellar Volume | Malformations of Cortical Development | Total |
| --- | --- | --- | --- | --- | --- | --- | --- | --- | --- | --- | --- | --- | --- | --- |
| Z002 | 2 | 2 | 0 | 2 | 2 | 2 | 3 | 0 | 0 | 1 | 0 | 0 | 3 | 17 |
| Z003 | 2 | 3 | 2 | 2 | 2 | 2 | 3 | 1 | 2 | 3 | 2 | 1 | 3 | 28 |
| Z004 | 3 | 3 | 2 | 3 | 3 | 3 | 3 | 3 | 3 | 4 | 1 | 1 | 3 | 35 |
| Z005 | 1 | 2 | 2 | 2 | 2 | 2 | 3 | 3 | 2 | 2 | 2 | 0 | 3 | 26 |
| Z006 | 2 | 2 | 2 | 2 | 2 | 2 | 3 | 3 | 1 | 3 | 1 | 0 | 3 | 26 |
| Z007 | 1 | 0 | 0 | 0 | 0 | 2 | 2 | 0 | 0 | 1 | 0 | 0 | 2 | 8 |
| Z008 | 3 | 3 | 2 | 2 | 2 | 3 | 3 | 1 | 3 | 3 | 1 | 0 | 3 | 29 |
| Z009 | 3 | 3 | 2 | 3 | 3 | 3 | 3 | 3 | 3 | 3 | 3 | 2 | 3 | 37 |
| Z010 | 2 | 3 | 1 | 3 | 3 | 3 | 3 | 3 | 3 | 3 | 3 | 2 | 3 | 35 |
| Z011 | 3 | 3 | 2 | 3 | 3 | 3 | 3 | 3 | 3 | 3 | 1 | 0 | 3 | 33 |
| Z012 | 3 | 3 | 2 | 3 | 3 | 3 | 3 | 3 | 3 | 3 | 3 | 3 | 3 | 38 |
| Z013 | 3 | 3 | 1 | 3 | 3 | 3 | 3 | 3 | 3 | 1 | 2 | 2 | 3 | 33 |
| Z014 | 2 | 3 | 2 | 2 | 2 | 2 | 3 | 3 | 3 | 2 | 2 | 1 | 3 | 30 |
| Z015 | 2 | 3 | 0 | 3 | 3 | 3 | 3 | 3 | 3 | 2 | 2 | 1 | 3 | 31 |
| Z016 | 2 | 2 | 1 | 2 | 2 | 2 | 3 | 1 | 3 | 2 | 2 | 2 | 3 | 27 |
| Z017 | 2 | 2 | 1 | 1 | 1 | 1 | 1 | 1 | 1 | 1 | 0 | 0 | 1 | 13 |
| Z018 | 3 | 3 | 2 | 3 | 3 | 3 | 3 | 3 | 3 | 2 | 2 | 1 | 3 | 34 |
| Z019 | 2 | 2 | 2 | 1 | 2 | 2 | 0 | 2 | 3 | 0 | 1 | 2 | 0 | 19 |
| Z020 | 1 | 1 | 1 | 1 | 2 | 0 | 0 | 2 | 3 | 0 | 2 | 0 | 0 | 13 |
| Z021 | 3 | 3 | 2 | 3 | 3 | 3 | 3 | 3 | 3 | 3 | 1 | 1 | 3 | 34 |
| Z022 | 2 | 2 | 2 | 1 | 2 | 3 | 3 | 0 | 2 | 2 | 0 | 0 | 3 | 22 |
| Z023 | 1 | 3 | 0 | 3 | 3 | 3 | 1 | 3 | 3 | 0 | 2 | 0 | 0 | 22 |
| Z024 | 0 | 0 | 0 | 0 | 1 | 2 | 0 | 0 | 1 | 1 | 0 | 0 | 0 | 5 |
| Z026 | 0 | 0 | 0 | 0 | 0 | 0 | 1 | 0 | 0 | 0 | 0 | 0 | 1 | 2 |
| Z027 | 3 | 3 | 0 | 3 | 3 | 2 | 3 | 1 | 3 | 3 | 2 | 0 | 3 | 29 |
| Z028 | 0 | 3 | 0 | 3 | 3 | 2 | 3 | 3 | 3 | 0 | 2 | 2 | 3 | 27 |
| Z029 | 3 | 3 | 0 | 3 | 3 | 3 | 3 | 3 | 3 | 3 | 2 | 1 | 3 | 33 |
| Z030 | 3 | 2 | 2 | 1 | 2 | 1 | 3 | 0 | 1 | 2 | 3 | 1 | 3 | 24 |
| Z101 | 2 | 3 | 0 | 3 | 3 | 3 | 0 | 2 | 1 | 0 | 0 | 0 | 0 | 17 |
| Z102 | 0 | 0 | 0 | 0 | 0 | 0 | 0 | 0 | 0 | 0 | 0 | 0 | 0 | 0 |
| Z103 | 1 | 2 | 1 | 2 | 2 | 2 | 3 | 3 | 2 | 2 | 3 | 3 | 3 | 29 |
| Z104 | 3 | 3 | 2 | 3 | 3 | 2 | 3 | 1 | 3 | 1 | 1 | 0 | 3 | 28 |
| Z105 | 2 | 2 | 1 | 2 | 2 | 2 | 3 | 1 | 2 | 2 | 0 | 0 | 3 | 22 |
| Z106 | 1 | 2 | 1 | 3 | 3 | 2 | 3 | 2 | 3 | 2 | 2 | 1 | 3 | 28 |
| Z107 | 1 | 0 | 0 | 0 | 1 | 1 | 0 | 0 | 0 | 1 | 0 | 0 | 0 | 4 |
| Z108 | 2 | 3 | 2 | 2 | 3 | 3 | 0 | 2 | 3 | 1 | 2 | 2 | 0 | 25 |
| Z109 | 0 | 0 | 0 | 0 | 0 | 0 | 1 | 0 | 0 | 1 | 0 | 0 | 1 | 3 |
| Z110 | 1 | 1 | 1 | 1 | 2 | 2 | 2 | 3 | 3 | 2 | 1 | 0 | 2 | 21 |
| Z111 | 1 | 1 | 1 | 1 | 1 | 1 | 0 | 1 | 1 | 0 | 1 | 1 | 0 | 10 |
| Z112 | 1 | 1 | 1 | 1 | 1 | 1 | 3 | 0 | 2 | 1 | 0 | 0 | 3 | 15 |
| Z113 | 1 | 1 | 1 | 2 | 2 | 1 | 3 | 0 | 1 | 0 | 0 | 0 | 3 | 15 |
| Z115 | 3 | 3 | 2 | 3 | 3 | 3 | 3 | 3 | 3 | 3 | 3 | 3 | 3 | 38 |
