## Supplementary material for "Zika Virus Congenital Microcephaly Severity Classification and the Association of Severity with Neuropsychomotor Development": Online supplementary material 8

**Online supplementary material 9**

Correlation table among Bayley scores, clinical measures, and severity classification.

|  | **Bayley Score** | | | | |
| --- | --- | --- | --- | --- | --- |
|  | **Cognition** | **Receptive Communication** | **Expressive Communication** | **Fine Motor** | **Gross Motor** |
| **HC at birth** | | | | | |
| **r** | 0.373 | 0.369 | 0.333 | 0.46 | 0.423 |
| **p-value** | 0.018 | 0.019 | 0.036 | 0.003 | 0.007 |
| **HC at the MRI exam** | | | | | |
| **r** | 0.21 | 0.109 | 0.274 | 0.298 | 0.303 |
| **p-value** | 0.273 | 0.575 | 0.15 | 0.117 | 0.11 |
| **HC growth ratio** | | | | | |
| **r** | -0.151 | -0.256 | -0.089 | -0.172 | -0.131 |
| **p-value** | 0.435 | 0.179 | 0.645 | 0.373 | 0.5 |
| **Zika virus-microcephaly severity score** | | | | | |
| **r** | -0.583 | -0.568 | -0.529 | -0.667 | -0.576 |
| **p-value** | < 0.001 | < 0.001 | < 0.001 | < 0.001 | < 0.001 |
| **Severity grouping based on Zika virus-microcephaly severity score** | | | | | |
| **r** | -0.538 | -0.577 | -0.561 | -0.629 | -0.6 |
| **p-value** | < 0.001 | < 0.001 | < 0.001 | < 0.001 | < 0.001 |
| **Semi-automated severity score** | | | | | |
| **r** | -0.609 | -0.597 | -0.542 | -0.707 | -0.628 |
| **p-value** | < 0.001 | < 0.001 | < 0.001 | < 0.001 | < 0.001 |
| **Severity grouping based on the semi-automated severity score** | | | | | |
| **r** | -0.478 | -0.501 | -0.444 | -0.562 | -0.492 |
| **p-value** | 0.002 | 0.001 | 0.004 | < 0.001 | 0.001 |

HC = Head circumference; r = Spearman partial correlation value (controlling for age).
