## Supplementary material for "Zika Virus Congenital Microcephaly Severity Classification and the Association of Severity with Neuropsychomotor Development": Online supplementary material 9

**Online supplementary material 10**

| **Participant** | **ZikV-microcephaly severity score** | **Semi-automated severity score** | **ZikV-microcephaly severity grouping** | **Semi-automated severity grouping** |
| --- | --- | --- | --- | --- |
| Z002 | 17 | 17 | Moderate | Moderate |
| Z003 | 28 | 24 | Severe | Moderate |
| Z004 | 35 | 32 | Severe | Severe |
| Z005 | 26 | 24 | Severe | Moderate |
| Z006 | 26 | 23 | Severe | Moderate |
| Z007 | 8 | 6 | Mild | Mild |
| Z008 | 29 | 29 | Severe | Severe |
| Z009 | 37 | 36 | Severe | Severe |
| Z010 | 35 | 35 | Severe | Severe |
| Z011 | 33 | 34 | Severe | Severe |
| Z012 | 38 | 36 | Severe | Severe |
| Z013 | 33 | 32 | Severe | Severe |
| Z014 | 30 | 25 | Severe | Moderate |
| Z015 | 31 | 31 | Severe | Severe |
| Z016 | 27 | 21 | Severe | Moderate |
| Z017 | 13 | 23 | Moderate | Moderate |
| Z018 | 34 | 30 | Severe | Severe |
| Z019 | 19 | 21 | Moderate | Moderate |
| Z020 | 13 | 13 | Moderate | Mild |
| Z021 | 34 | 34 | Severe | Severe |
| Z022 | 22 | 18 | Moderate | Moderate |
| Z023 | 22 | 27 | Moderate | Moderate |
| Z024 | 5 | 3 | Mild | Mild |
| Z026 | 2 | 1 | Mild | Mild |
| Z027 | 29 | 25 | Severe | Moderate |
| Z028 | 27 | 25 | Severe | Moderate |
| Z029 | 33 | 32 | Severe | Severe |
| Z030 | 24 | 31 | Moderate | Severe |
| Z101 | 17 | 30 | Moderate | Severe |
| Z102 | 0 | 3 | Mild | Mild |
| Z103 | 29 | 25 | Severe | Moderate |
| Z104 | 28 | 28 | Severe | Severe |
| Z105 | 22 | 21 | Moderate | Moderate |
| Z106 | 28 | 26 | Severe | Moderate |
| Z107 | 4 | 8 | Mild | Mild |
| Z108 | 25 | 31 | Moderate | Severe |
| Z109 | 3 | 8 | Mild | Mild |
| Z110 | 21 | 25 | Moderate | Moderate |
| Z111 | 10 | 12 | Mild | Mild |
| Z112 | 15 | 11 | Moderate | Mild |
| Z113 | 15 | 17 | Moderate | Moderate |
| Z115 | 38 | 36 | Severe | Severe |
